# Circulating insulin-like growth factor-I and risk of 25 common conditions: outcome-wide analyses in the UK Biobank Study

**DOI:** 10.1101/2021.05.05.21256665

**Authors:** Keren Papier, Anika Knuppel, Aurora Perez-Cornago, Eleanor L. Watts, Tammy YN Tong, Julie A Schmidt, Naomi Allen, Timothy J Key, Ruth C Travis

## Abstract

**Background:** While there is strong epidemiological evidence that circulating insulin-like growth factor-I (IGF-I) is associated with a higher risk of several cancers, little is known about its association with non-cancer outcomes. We investigated associations of circulating IGF-I with risk of 25 common conditions, other than cancer, in a large British cohort.

**Methods:** Study participants were 318 749 middle-aged adults enrolled in the UK Biobank Study. Serum IGF-I concentration was measured in samples collected at baseline (2006-2010), and re-measured in 12 334 participants after an average of 4.3 years. We followed-up participants over an average of 11.5 years by linking to hospital admissions and mortality registries. Multivariable-adjusted Cox regressions estimated hazard ratios (HRs) and 95% confidence intervals (CIs) for associations between circulating IGF-I and 25 common conditions, using the repeated IGF-I measurements to correct for regression dilution bias.

**Results:** After correction for multiple testing (*P*<0.002), IGF-I was positively associated with carpal tunnel syndrome (HR per 5 nmol/l higher concentration=1.12, 95% CI, 1.08-1.16), and inversely associated with varicose veins (0.90, 0.85-0.95), cataracts (0.97, 0.95-0.99), diabetes (0.92, 0.90-0.95), and iron deficiency anaemia (0.90, 0.86-0.93). The associations for cataracts and diabetes attenuated when restricted to cases diagnosed after five or more years of follow-up, suggesting that these associations were likely affected by reverse causality.

**Conclusions:** Higher IGF-I concentration might be associated with the risk for several conditions, but genetic studies are needed to clarify which associations may be causal.

**Key messages:**

- Evidence on the association between circulating insulin-like growth factor-I (IGF-I) and risk of many common, non-cancer health outcomes in the general population is relatively limited.
- This study used an outcome-wide approach to prospectively examine associations of circulating IGF-I with risk of 25 common conditions in a large, prospective cohort of over 300,000 UK adults.
- Our study showed that circulating IGF-I is associated with risks of several common diseases and conditions; IGF-I was positively associated with carpal tunnel syndrome and inversely associated with varicose veins, cataracts, diabetes, and iron deficiency anaemia.
- Further research is needed to evaluate whether these differences in risk may reflect causal relationships.

## Background

Insulin-like growth factor-I (IGF-I) is a polypeptide hormone that is primarily synthesised in the liver following growth hormone stimulation,^1^ and promotes tissue growth and development in multiple organ systems by acting as a primary mediator of the effects of growth hormone.^2^ Clinically high circulating IGF-I concentration, as in adults with acromegaly, is associated with a higher risk of several diseases,^3^ particularly higher risks of cardiovascular disease, metabolic disorders (e.g. insulin resistance), biliary diseases (e.g. gallbladder disease), gastrointestinal diseases (e.g. colon polyps), arthropathy, musculoskeletal disorders (e.g. carpal tunnel syndrome), genitourinary diseases (e.g. kidney stones, enlarged prostate^4^), respiratory disease, sleep apnoea, and some cancers.^5,6^

Higher IGF-I concentrations in adults without acromegaly have been shown to also be associated with increased risks of several cancers,^7^ but corresponding evidence for non-cancer outcomes is inconsistent and/or limited to cross-sectional design. Although some of the available prospective observational and genetic evidence suggests that higher IGF-I levels might be positively associated with type 2 diabetes,^8,9^ ischaemic heart disease (IHD),^9-11^ hip and knee osteoarthritis,^12^ enlarged prostate,^13^ and colon adenomas,^14,15^ some studies also reported null,^16-24^ and inverse ^25^ associations for these outcomes. These equivocal findings and the lack of available prospective evidence for many non-cancer outcomes likely relates at least in part to outcome-selection bias, small sample sizes, and/or the relatively short follow-up of previous studies. To address this, we used an outcome-wide approach to examine the prospective association of circulating IGF-I with 25 common, non-cancerous diseases and conditions, in a large British study of men and women with over 10 years of follow-up.

## Methods

### Study population

The UK Biobank is a prospective cohort study of middle-aged men and women (aged 40-69 years at recruitment) from the general population across the UK.^26^ Approximately 9.2 million individuals who were registered with the National Health Service (NHS) and lived within about 40 km of one of 22 UK Biobank assessment centres were invited to participate, of whom 503 317 (5.5%) joined the cohort between 2006 and 2010.^27^ The UK Biobank study was approved by the North West Multi-Centre Research Ethics Committee (reference number 16/NW/0274).

### Laboratory assessment

All participants provided a non-fasting blood sample at recruitment and approximately 20 000 participants (21% of those re-invited) agreed to provide an additional blood sample as part of a repeat assessment between 2012 and 2013 (https://biobank.ctsu.ox.ac.uk/~bbdatan/Repeat_assessment_doc_v1.0.pdf). IGF-I concentration was measured in serum samples which had been stored at −80°C, using chemiluminescent immunoassays (DiaSorin Liaison XL, analytical range 1.3-195 nmol/l). ^28^ The average within-laboratory coefficients of variation (ratio of the standard deviation to the mean for quality control samples) ranged from 5.3-6.2%. Full details of the assay methods and quality assurance protocols are available online (https://biobank.ndph.ox.ac.uk/showcase/showcase/docs/serum_biochemistry.pdf).

### Assessment of other characteristics

Participants provided information on their personal, physical, sociodemographic and other lifestyle characteristics at the baseline assessment visit via a self-completed touchscreen questionnaire and a computer assisted personal interview. Participants also underwent physical measurements (e.g. weight and height) during the recruitment visit.

### Assessment of health outcomes

Participants’ health was followed up via record linkage to routine health records, including national death and cancer registers and in-patient hospital admissions. Outcomes of interest were the 25 most common, well-defined primary causes of non-cancer related hospital admission in this cohort based on the primary International Classification of Diseases (ICD) 10 diagnosis codes recorded during admission.^29^ We excluded some common reasons for hospital admission in this cohort (e.g. nausea or heartburn) because they were not well-defined and/or were likely to be associated with a diverse range of underlying conditions. We included diabetes even though it was not among the 25 most common primary diagnoses associated with admission, as it is a common secondary (i.e. co-incident or underlying) reason for admission (See Supplementary Methods M1 and Supplementary Table S1 for information on censor dates, diagnosis, and procedure codes.)

### Exclusions

Of the 503 317 participants recruited, 829 were excluded because they had withdrawn from the study, and 35 463 were excluded because they had missing data on any biomarkers and IGF-I concentration at baseline. We additionally excluded participants whose genetic sex differed from their reported gender (n=355), those with missing data for weight (n=1553) or height (n=307), prevalent cancer (except non-melanoma skin cancer, ICD-10 C44) prior to recruitment (n=24 626), those who did not self-report good or excellent health (n=110 830), those with prevalent diabetes or unknown diabetes status (n=10 425), and those with less than one year of follow-up (n=180) to reduce the risk of reverse causality, resulting in study sample of 318 749 participants. (Supplementary Figure S1).

### Statistical analysis

We used Cox proportional hazards regression models, with age as the underlying time variable, to estimate hazard ratios (HRs) and 95% confidence intervals (CIs) for associations between IGF-I concentration and each condition of interest, with Bonferroni correction to allow for multiple testing (*P*<0.002 for 25 tests). IGF-I concentrations were modelled both continuously (per 5 nmol/l of IGF-I concentration, equivalent to ∼1 SD in the cohort) and categorically (sex-specific fifths). Each endpoint was assessed using a separate analysis with participants contributing person-years at-risk for each condition of interest for that analysis. We calculated person-years of follow-up using participants’ age at recruitment and their age at hospital admission, death, or loss or end of follow up (November 30^th^, 2020 for England, October 31^st^, 2020 for Scotland, and February 28^th^, 2018 for Wales).

The use of a single measure can lead to under-estimation of risks due to within-person variability and laboratory measurement error (“regression dilution bias”).^30^ Therefore, we used the repeated IGF-I measures collected from 12 334 participants (who met our inclusion criteria) on average 4.3 years (SD 0.9 years) after recruitment to correct HRs for trend using the McMahon-Peto method;^31^ the log HRs and standard errors were divided by the sex-specific regression dilution ratios from the subsample, which were obtained by dividing the difference in the mean IGF-I concentrations between the 5^th^ and 1^st^ fifths in the repeat measure by the equivalent mean difference in the baseline measure.

All analyses were stratified by age group (<45, 45-49, 50-54, 55-59, 60-64, ≥65 years), sex and geographical region (London, North-West, North-East, Yorkshire and Humber, West Midlands, East Midlands, South-East, South-West, Wales, and Scotland) (Model 0). In Model 1, we additionally adjusted for ethnicity (White, non-White, unknown), socio-economic deprivation (Townsend index quintiles, unknown), qualification (college or university degree/vocational qualification, national examination at ages 17-18 years, national examination at age 16 years, unknown), smoking (never, former, current <15 cigarettes/day, current >15 cigarettes/ day, unknown), physical activity (<10, 10-19, 20-39, 40-59, >60 MET hours per week, unknown), alcohol intake (<1.0, 1.0–4.9, 5.0–9.9, 10.0–14.9, 15.0–19.9, 20.0–24.9, and ≥25.0 g/day, non-drinker, and unknown), and height (continuous). For women, Model 1 was additionally adjusted for menopausal status (pre-, postmenopausal, unknown), hormone-replacement therapy (HRT) use (never, past, current, unknown), oral contraceptive pill (OCP) use (never, past, current, unknown), and parity (nulliparous, 1-2, 3 or more, unknown). For Model 2, we further adjusted for body mass index (BMI) (<20.0, 20.0-22.49, 22.5-24.99, 25.0-27.49, 27.5-29.99, 30.0-32.49, 32.5-34.99, >35.0 kg/m^2^). An ‘unknown’ category was created for each covariate with missing data (proportion of missings ranged from <1% for alcohol to 20% for physical activity).

### Sensitivity analyses

We assessed the potential for residual confounding by other relevant biomarkers previously found to be associated with IGF-I and some of the outcomes^32,33^ by additionally adjusting our main model (Model 2) for serum concentrations of C-reactive protein, glycated haemoglobin, and sex hormone–binding globulin (fifths, unknown for each) (Model 3). We also assessed heterogeneity by follow-up time at diagnosis (less than five years and five years or over) using stratified Cox regression models, comparing HRs and standard errors in the two subgroups using a χ^2^ test for heterogeneity (*P*<0.05).

All analyses were conducted using STATA version 16.1 (Stata Corp LP, College Station, TX, USA). All *P* values were two-sided.

## Results

### Baseline characteristics

Table 1 presents the cohort characteristics of study participants by mean baseline levels of circulating IGF-I. We observed higher IGF-I concentrations in men, younger adults, those who self-identified to be of non-White ethnicity, those in the top fifth of height, adults with a BMI in the middle range (between 22.5 and 27.5 kg/m^2^) compared to those with a lower and higher BMI, adults who were affluent and had a higher level of attained education, non-smokers, moderate alcohol consumers, adults with low levels of physical activity, and women who had never used hormone replacement therapy, were using the oral contraceptive pill, did not have children, and were premenopausal. At baseline, the mean circulating IGF-I concentration was 21.7 nmol/l (SD 5.5). The correlation coefficient between IGF-I measured at baseline and at follow-up was 0.77.

**Table 1.**
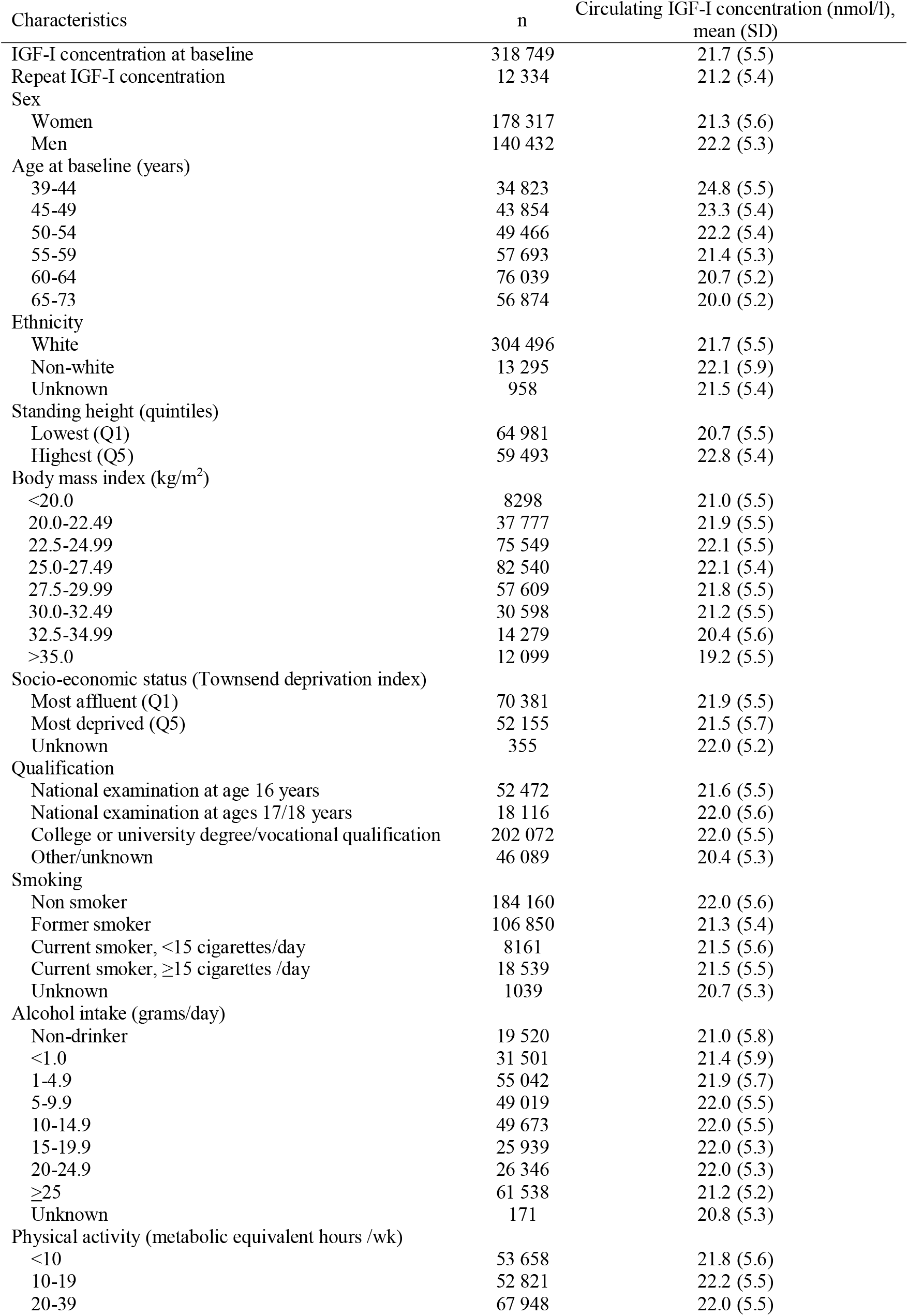
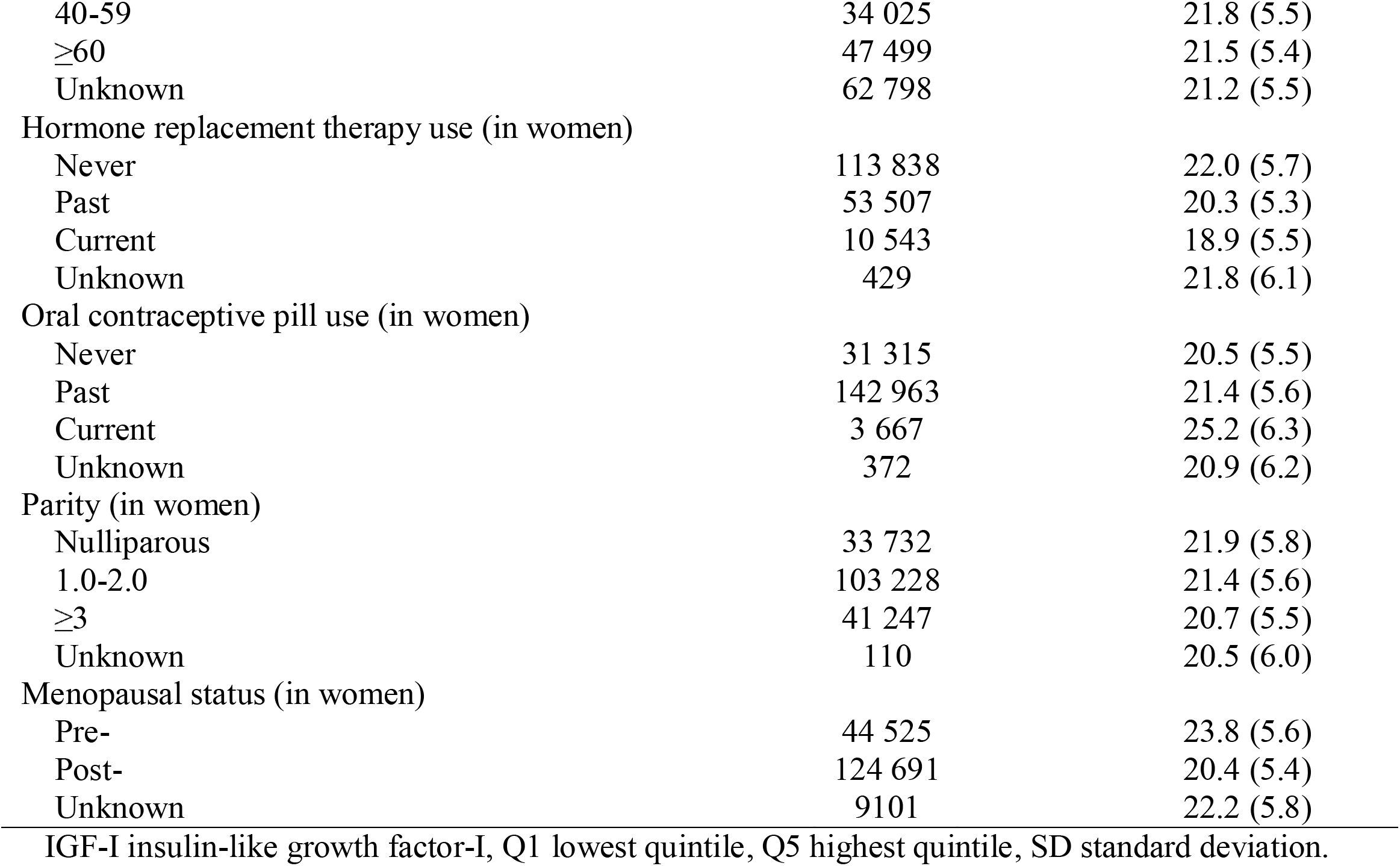
Baseline characteristics by serum IGF-I levels in UK Biobank participants (n=318 749).

| Characteristics | n | Circulating IGF-I concentration (nmol/l),<br>mean (SD) |
| --- | --- | --- |
| IGF-I concentration at baseline | 318 749 | 21.7 (5.5) |
| Repeat IGF-I concentration | 12 334 | 21.2 (5.4) |
| Sex |  |  |
| Women | 178 317 | 21.3 (5.6) |
| Men | 140 432 | 22.2 (5.3) |
| Age at baseline (years) |  |  |
| 39-44 | 34 823 | 24.8 (5.5) |
| 45-49 | 43 854 | 23.3 (5.4) |
| 50-54 | 49 466 | 22.2 (5.4) |
| 55-59 | 57 693 | 21.4 (5.3) |
| 60-64 | 76 039 | 20.7 (5.2) |
| 65-73 | 56 874 | 20.0 (5.2) |
| Ethnicity |  |  |
| White | 304 496 | 21.7 (5.5) |
| Non-white | 13 295 | 22.1 (5.9) |
| Unknown | 958 | 21.5 (5.4) |
| Standing height (quintiles) |  |  |
| Lowest (Q1) | 64 981 | 20.7 (5.5) |
| Highest (Q5) | 59 493 | 22.8 (5.4) |
| Body mass index (kg/m<sup>2</sup>) |  |  |
| <20.0 | 8298 | 21.0 (5.5) |
| 20.0-22.49 | 37 777 | 21.9 (5.5) |
| 22.5-24.99 | 75 549 | 22.1 (5.5) |
| 25.0-27.49 | 82 540 | 22.1 (5.4) |
| 27.5-29.99 | 57 609 | 21.8 (5.5) |
| 30.0-32.49 | 30 598 | 21.2 (5.5) |
| 32.5-34.99 | 14 279 | 20.4 (5.6) |
| >35.0 | 12 099 | 19.2 (5.5) |
| Socio-economic status (Townsend deprivation index) |  |  |
| Most affluent (Q1) | 70 381 | 21.9 (5.5) |
| Most deprived (Q5) | 52 155 | 21.5 (5.7) |
| Unknown | 355 | 22.0 (5.2) |
| Qualification |  |  |
| National examination at age 16 years | 52 472 | 21.6 (5.5) |
| National examination at ages 17/18 years | 18 116 | 22.0 (5.6) |
| College or university degree/vocational qualification | 202 072 | 22.0 (5.5) |
| Other/unknown | 46 089 | 20.4 (5.3) |
| Smoking |  |  |
| Non smoker | 184 160 | 22.0 (5.6) |
| Former smoker | 106 850 | 21.3 (5.4) |
| Current smoker, <15 cigarettes/day | 8161 | 21.5 (5.6) |
| Current smoker, ≥15 cigarettes /day | 18 539 | 21.5 (5.5) |
| Unknown | 1039 | 20.7 (5.3) |
| Alcohol intake (grams/day) |  |  |
| Non-drinker | 19 520 | 21.0 (5.8) |
| <1.0 | 31 501 | 21.4 (5.9) |
| 1-4.9 | 55 042 | 21.9 (5.7) |
| 5-9.9 | 49 019 | 22.0 (5.5) |
| 10-14.9 | 49 673 | 22.0 (5.5) |
| 15-19.9 | 25 939 | 22.0 (5.3) |
| 20-24.9 | 26 346 | 22.0 (5.3) |
| ≥25 | 61 538 | 21.2 (5.2) |
| Unknown | 171 | 20.8 (5.3) |
| Physical activity (metabolic equivalent hours /wk) |  |  |
| <10 | 53 658 | 21.8 (5.6) |
| 10-19 | 52 821 | 22.2 (5.5) |
| 20-39 | 67 948 | 22.0 (5.5) |
| 40-59 | 34 025 | 21.8 (5.5) |
| ≥60 | 47 499 | 21.5 (5.4) |
| Unknown | 62 798 | 21.2 (5.5) |
| Hormone replacement therapy use (in women) |  |  |
| Never | 113 838 | 22.0 (5.7) |
| Past | 53 507 | 20.3 (5.3) |
| Current | 10 543 | 18.9 (5.5) |
| Unknown | 429 | 21.8 (6.1) |
| Oral contraceptive pill use (in women) |  |  |
| Never | 31 315 | 20.5 (5.5) |
| Past | 142 963 | 21.4 (5.6) |
| Current | 3 667 | 25.2 (6.3) |
| Unknown | 372 | 20.9 (6.2) |
| Parity (in women) |  |  |
| Nulliparous | 33 732 | 21.9 (5.8) |
| 1.0-2.0 | 103 228 | 21.4 (5.6) |
| ≥3 | 41 247 | 20.7 (5.5) |
| Unknown | 110 | 20.5 (6.0) |
| Menopausal status (in women) |  |  |
| Pre- | 44 525 | 23.8 (5.6) |
| Post- | 124 691 | 20.4 (5.4) |
| Unknown | 9101 | 22.2 (5.8) |

IGF-I insulin-like growth factor-I, Q1 lowest quintile, Q5 highest quintile, SD standard deviation.

### Risk analyses

Figure 1 presents the HRs and 95% CIs for 25 common conditions in relation to a per 5 nmol/l higher serum IGF-I concentration, ordered by disease subgroup (circulatory, respiratory, digestive, joint disorder, genitourinary and other diseases) and effect size; estimated using multivariable-adjusted Cox regression models (Model 2), corrected for regression dilution bias. After correction for multiple testing (*P*<0.002), there was a positive association between IGF-I concentration and carpal tunnel syndrome (HR per 5 nmol/l higher concentration=1.12, 95% CI, 1.08-1.16), and inverse associations with varicose veins (0.90, 0.85-0.95), cataracts (0.97, 0.95-0.99), diabetes (0.92, 0.90-0.95), and iron deficiency anaemia (0.90, 0.86-0.93). Similar associations were observed by sex-specific fifths of IGF-I concentration for these outcomes: carpal tunnel syndrome (HR in the top versus the lowest fifth=1.34, 95% CI, 1.20-1.50), varicose veins (0.75, 0.64-0.87), cataracts (0.91, 0.87-0.95), diabetes (0.85, 0.79-0.91), and iron deficiency anaemia (0.77, 0.69-0.85) (Supplementary Table S2). Higher IGF-I concentration was also associated with lower risks of atrial fibrillation and flutter, pneumonia, gastritis and duodenitis, and noninfective enteritis and colitis, and higher risks of haemorrhoids, osteoarthritis, and enlarged prostate at the *P*<0.05 level.

**Figure 1.**
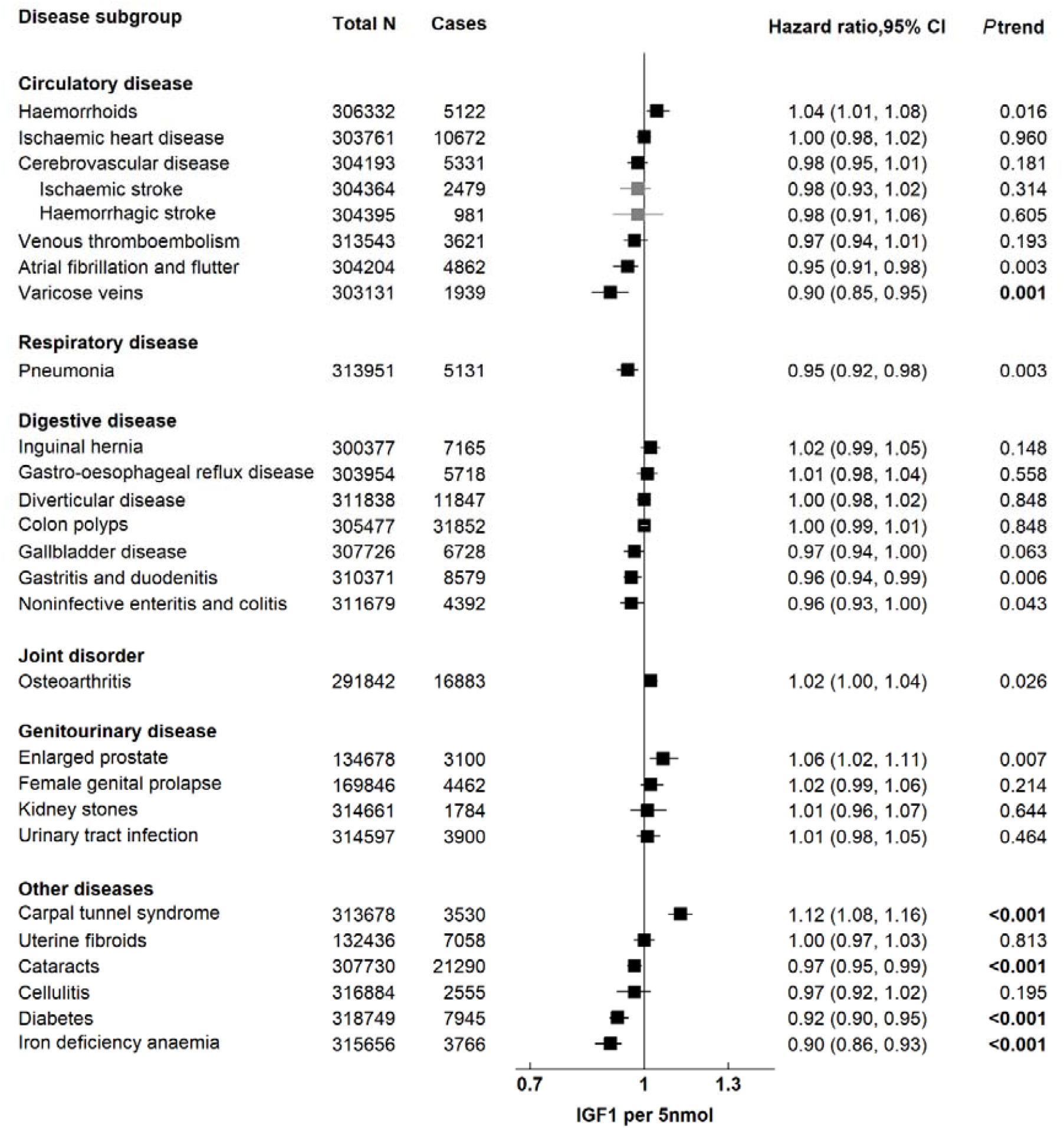
Relative risk of 25 common conditions per 5 nmol/l higher IGF-I concentration in UK Biobank, corrected for regression dilution bias. Stratified for age group (<45, 45–49, 50–54, 55–59, 60–64, and ≥65 years), sex and region and adjusted for age (underlying time variable), ethnicity (White, non-white, unknown), deprivation (Townsend index quintiles, unknown), qualification (College or university degree/vocational qualification, National examination at ages 17-18,National examination at age 16, unknown), smoking (never, former, current <15 cigarettes/day, current >15 cigarettes/ day, unknown), physical activity (<10, 10-<20, 20-40, 40-<60, >=60 MET hours per week, unknown), alcohol intake (<1.0, 1.0–4.9, 5.0–9.9, 10.0–14.9, 15.0–19.9, 20.0–24.9, and ≥25.0 g/day, non-drinker, and unknown), height (continuous), and in women: menopausal status (pre-, postmenopausal, unknown), hormone-replacement therapy (never, past, current, unknown), oral contraceptive pill intake (never, past, current, unknown), parity (nulliparous, 1-2, 3 or more, unknown), and body mass index (<20.0, 20.0-22.49, 22.5-24.99, 25.0-27.49, 27.5-29.99, 30.0-32.49, 32.5-34.99, >35.0). CI confidence intervals, IGF-I insulin-like growth factor-I. *P* trend in bold: *P* value statistically significant after Bonferroni correction (*P*<0.002).

### Sensitivity analyses

We observed similar results to those from the main analysis in sensitivity analyses adjusting for serum concentrations of C-reactive protein, glycated haemoglobin, and sex hormone– binding globulin (Supplementary Table S2, Model 3).

Figure 2 shows associations between risk of 25 common conditions and higher IGF-I concentration (per 5 nmol/l) stratified by follow-up time at diagnosis (less than five years and five years or over). There was evidence of heterogeneity by follow-up time for associations with cataracts (Phet=0.040) and diabetes (Phet=0.008), with associations closer to null in participants diagnosed after five or more years of follow-up. We also observed positive associations for IHD, haemorrhoids, colon polyps, osteoarthritis, kidney stones, and uterine fibroids, and an inverse association for pneumonia in participants diagnosed after five or more years of follow-up (Phet<0.05).

**Figure 2.**
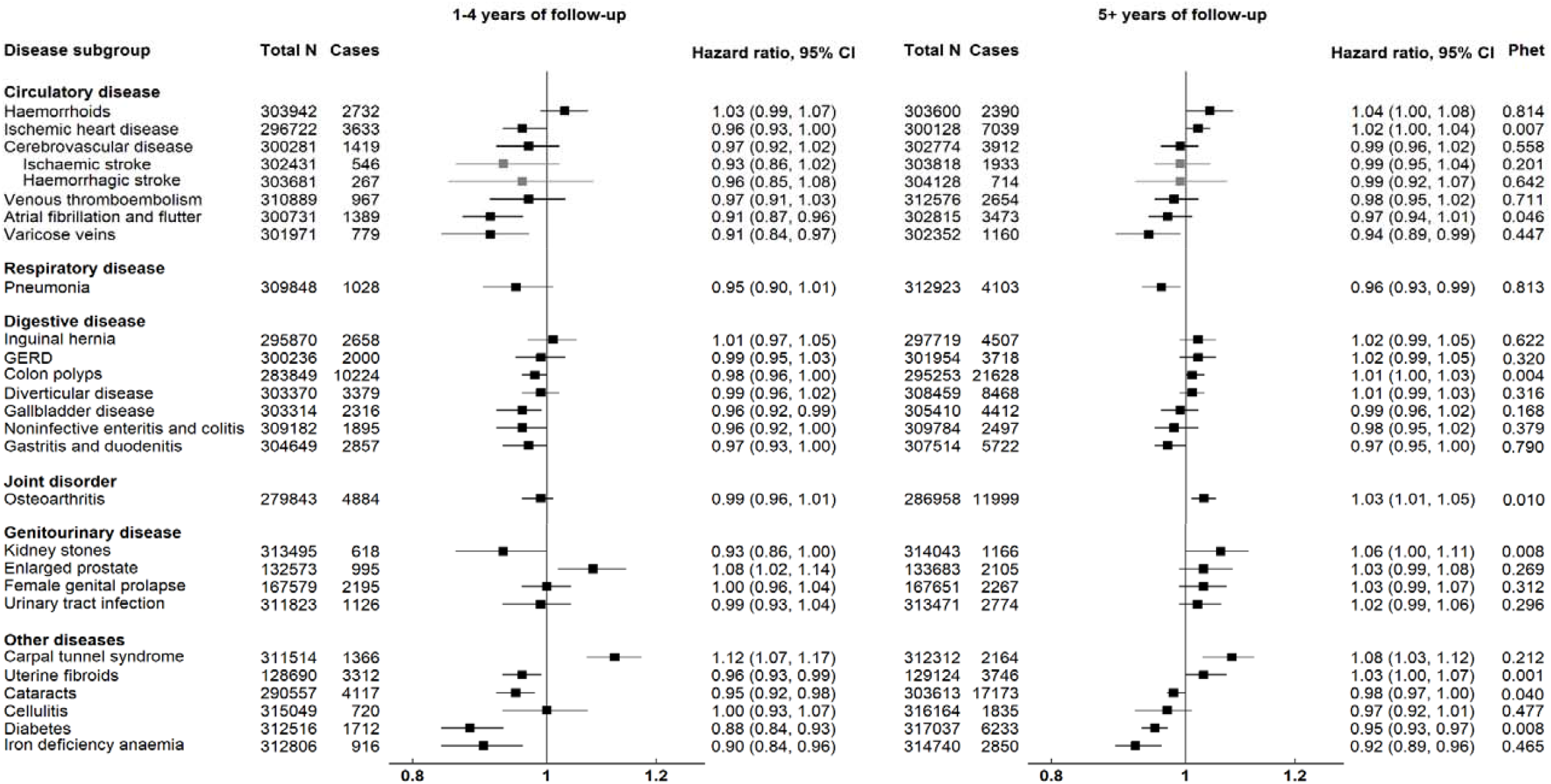
Relative risk of 25 common conditions per 5 nmol/l higher IGF-I concentration by follow-up time at diagnosis. Stratified for age group (<45, 45–49, 50–54, 55–59, 60–64, and ≥65 years), sex and region and adjusted for age (underlying time variable), ethnicity (White, non-white, unknown), deprivation (Townsend index quintiles, unknown), qualification (College or university degree/vocational qualification, National examination at ages 17-18,National examination at age 16, unknown), smoking (never, former, current <15 cigarettes/day, current >15 cigarettes/ day, unknown), physical activity (<10, 10-<20, 20-40, 40-<60, >=60 MET hours per week, unknown), alcohol intake (<1.0, 1.0–4.9, 5.0–9.9, 10.0–14.9, 15.0–19.9, 20.0–24.9, and ≥25.0 g/day, non-drinker, and unknown), height (continuous), and in women: menopausal status (pre-, postmenopausal, unknown), hormone-replacement therapy (never, past, current, unknown), oral contraceptive pill intake (never, past, current, unknown), parity (nulliparous, 1-2, 3 or more, unknown), and body mass index (<20.0, 20.0-22.49, 22.5-24.99, 25.0-27.49, 27.5-29.99, 30.0-32.49, 32.5-34.99, >35.0). GERD Gastro-oesophageal reflux disease, CI confidence intervals, IGF-I insulin-like growth factor-I.

## Discussion

In this large prospective and outcome-wide investigation of associations between circulating IGF-I and a range of non-cancer outcomes, we found that, after accounting for multiple testing and regression dilution bias, higher IGF-I concentration was positively associated with risk of carpal tunnel syndrome and inversely associated with incident varicose veins, cataracts, diabetes, and iron deficiency anaemia. The associations for carpal tunnel syndrome, varicose veins, and iron deficiency anaemia did not vary by follow-up time at diagnosis. For cataracts and diabetes though, the associations were closer to null in those diagnosed after five or more years of follow-up, suggesting that these associations might have been affected by reverse causality (whereby IGF-I levels change as a result of early pathophysiological processes).

To our knowledge, this is the first study of IGF-I and risk of carpal tunnel syndrome in a general, population-based cohort. In line with our results, one case-control study found that 34 adults with acromegaly had larger peripheral nerves, and that biochemical control of IGF-I concentrations over a one-year follow-up resulted in reduced nerve size.^34^ The positive association between circulating IGF-I and carpal tunnel syndrome could be plausible due to IGF-I’s involvement in nerve growth and formation;^35^ in adults with acromegaly, carpal tunnel syndrome has been attributed to median nerve enlargement, which was correlated with circulating IGF-I.^36,37^

We found a small inverse association between IGF-I and risk of varicose veins. This is a novel epidemiological finding, although an *in-vitro* study found that IGF-I was associated with the proliferation of smooth muscle cells of saphenous veins^38^. IGF-I’s role in promoting the growth and survival of smooth muscle cells would suggest that the inverse association we observed is plausible.

We also found an inverse association between IGF-I concentration and iron deficiency anaemia. We are not aware of any published prospective evidence, but previous cross-sectional studies have found that low IGF-I concentrations were associated with lower haemoglobin concentration and higher prevalence of anaemia in middle-aged and elderly adults.^39-42^ IGF-I might play an important role in erythropoiesis,^43^ suggesting that the observed inverse association is plausible. However, iron deficiency anaemia might have a longer lag time from disease onset or initial diagnosis to hospital admission,^44^ therefore reverse causality cannot be ruled out and longer follow-up is needed.

We found a novel inverse association between IGF-I and risk of cataracts. However, in analyses stratified by follow-up time at diagnosis this appeared to be due to reverse causality, possibly related to a shared pathophysiology with insulin resistance. In support of our findings, in adults with acromegaly, visual disturbances appear to relate to the effects of space-occupying lesions of pituitary adenomas rather than to circulating IGF-I levels.^45^ Also, some evidence from *in-vitro* rat models has shown that IGF-I might decrease the amount of α-crystallin (a lens protein) made in the lens fibre cells^46^, therefore this might be expected to lead to a positive association of IGF-I with risk of cataracts since higher levels of α-crystallin have been associated with a lower risk of cataract formation.^46^

Recent prospective and genetic evidence has suggested that there could be a positive association between circulating IGF-I concentration and type 2 diabetes risk,^8,9,16^ possibly due to its involvement in glucose homeostasis.^47^ This is in contrast to the inverse association we observed for incident diabetes (which is likely to be mostly type 2 diabetes due to the older age structure of this cohort). However, our findings may be due to reverse causality. Diabetes may go undiagnosed for years,^48^ therefore a long follow-up period is needed to avoid picking-up prevalent cases or pre-clinical disease.

We also found evidence for positive associations between IGF-I and IHD, haemorrhoids, colon polyps, osteoarthritis, kidney stones, and uterine fibroids, and an inverse association with pneumonia in participants diagnosed after five or more years of follow-up. The findings for IHD,^9^ osteoarthritis,^12^ colon polyps,^14,15^ and uterine fibroids^49^ are in line with some of the available prospective or genetic evidence from population-based studies, and the results for kidney stones are supported by studies in adults with acromegaly.^5^ It is possible that some of these associations were masked by reverse causality in the first five years of follow-up, and follow-up with larger numbers is needed to clarify whether IGF-I might associate with these outcomes.

This is the first study to adopt an outcome-wide approach to the investigation of IGF-I and risk of 25 common conditions (other than cancer); this comprehensive approach allowed us to assess and compare the effect sizes of multiple outcomes within the UK Biobank and reduce outcome-selection bias. Additional strengths of our study include its population-based design, the use of national record linkage to ascertain information on disease incidence, which eliminates misclassification and reduces attrition bias at follow-up, and its large size; this is the largest prospective study of IGF-I and most of these 25 common conditions.

Nevertheless, our study is not without its limitations. Some measurement error could have occurred when measuring IGF-I at baseline, but we reduced the potential impact of regression dilution bias by correcting baseline measures in all participants with a repeat IGF-I measure from a subsample of participants. Furthermore, we cannot rule-out that multiple testing could have led to some chance findings, though we addressed this using Bonferroni correction. However, this is a strict approach and it is possible that some of the associations with a *P*<0.05 do not reflect a chance finding. Additionally, because we used an outcome-wide approach, it is possible that we did not fully adjust for confounders that might have affected some of the individual conditions. However, many of these associations are exploratory and therefore not all of the confounders are known. Moreover, some conditions might go undiagnosed for some time and only require hospital care at later stages, and therefore might reflect prevalent or preclinical disease and/or more severe disease (such as in the case of diabetes). Furthermore, IGF-I–related proteins such as IGF-II and IGF-binding proteins (IGFBP), which play a role in the regulation of IGF-I bioavailability and signalling,^50^ were not measured in this study. Therefore, the observed associations could partially reflect other aspects of the IGF signalling pathway. Finally, the UK Biobank is predominantly made up of white Europeans, so the generalisability of our findings might be limited.

## Conclusions

The findings from this large population-based study suggest that circulating IGF-I may be associated with several common conditions (other than cancer), including positively with risk of carpal tunnel syndrome and inversely with risks of varicose veins and iron deficiency anaemia. Additional studies, including genetic analyses, are needed to assess whether these findings reflect casual relationships.

## Data Availability

All bona fide researchers can apply to use the UK Biobank resource for health related research that is in the public interest (https://www.ukbiobank.ac.uk/register-apply/).

## Acknowledgements

This research has been conducted using the UK Biobank Resource under application number 24494. All bona fide researchers can apply to use the UK Biobank resource for health related research that is in the public interest (https://www.ukbiobank.ac.uk/register-apply/). We thank Georgina K Fensom for her valuable statistical advice and all of the participants, researchers and support staff who make the study possible.

## Authors’ contributions

The research question was conceived and designed by KP, TJK, NA and RCT. KP analysed the data, prepared the figures and tables, and wrote the first draft of the manuscript; KP and AK prepared the data for analysis; and AK, AP-C, EW, TYNT, JS, NA, TJK, and RCT, provided input on data analysis and interpretation of results. All authors revised the manuscript critically for important intellectual content, and read and approved the final manuscript.

