## Supplemental material for "Circulating insulin-like growth factor-I and risk of 25 common conditions: outcome-wide analyses in the UK Biobank Study"

### Supplementary Methods

#### Supplementary Methods M1. Assessment of health outcomes.

For participants in England, Hospital Episode Statistics (HES) and information on date and cause of death were available until November 30^th^, 2020; for participants in Scotland, the Scottish Morbidity Records and information on date and cause of death were available until October 31^st^, 2020; and for participants in Wales, the Patient Episode Database and information on date and cause of death were available until February 28^th^, 2018. We also obtained information on cancer registrations (including date and cancer site) from the NHS Central Registers. <https://biobank.ndph.ox.ac.uk/showcase/exinfo.cgi?src=Data_providers_and_dates>

Disease end points, information on diagnoses or procedures associated with hospital admissions, and causes of death were all coded according to the 9^th^ or 10^th^ revisions of the World Health Organization’s International Classification of Diseases (ICD-9 and ICD-10), and the Office of Population Censuses and Surveys classification of surgical operations and procedures (OPCS-4), fourth revision. (See Supplementary Table S1 for information on exclusion, diagnosis and procedure codes).

### Supplementary Tables

| Supplementary Table S1. Disease outcome definition and exclusion criteria. | | | | | | | | |
| --- | --- | --- | --- | --- | --- | --- | --- | --- |
| Admission cause | Outcome definition using ICD-10 | Relevant procedure code using OPSC-4 definition | Exclusion criteria using ICD-10 | Exclusion criteria using OPSC-4 | Exclusion criteria using ICD-9 | Exclusion criteria using touchscreen | Exclusion criteria using UKB interviews for outcomes^a^ | Exclusion criteria using UKB interviews for procedures^b^ |
| Circulatory disease |  |  |  |  |  |  |  |  |
| Ischaemic heart diseases | I20, I21, I22, I23, I24, I25 |  | I48, I20, I21, I22, I23, I24, I25, G45, I60, I61, I62, I63, I64, I65, I66, I67, I68, I69 |  | 427.31, 427.32, 410, 411, 412, 413, 414, 430, 431, 432, 433, 434, 435, 436, 437, 438 | UKB variable 'Has a doctor ever told you that you have the following conditions 6150?'  1) Heart attack 2) angina  3) stroke | 1471 atrial fibrillation  1483 atrial flutter  1074 angina  1075 heart attack/myocardial infarction  1081 stroke  1082 transient ischaemic attack1083 subdural haemorrhage/haematoma  1086 subarachnoid haemorrhage |  |
| Atrial fibrillation and flutter | I48 |  | I48, I20, I21, I22, I23, I24, I25, G45, I60, I61, I62, I63, I64, I65, I66, I67, I68, I69 |  | 427.31, 427.32, 410, 411, 412, 413, 414, 430, 431, 432, 433, 434, 435, 436, 437, 438 | UKB variable 'Has a doctor ever told you that you have the following conditions 6150?'  1) Heart attack, 2) angina,  3) stroke | 1471 atrial fibrillation  1483 atrial flutter  1074 angina  1075 heart attack/myocardial infarction  1081 stroke  1082 transient ischaemic attack1083 subdural haemorrhage/haematoma  1086 subarachnoid haemorrhage |  |
| Cerebrovascular disease | G45 I60 I61 I62 I63 I64 I65 I66 I67 I68 I69 |  | I48, I20, I21, I22, I23, I24, I25, G45, I60, I61, I62, I63, I64, I65, I66, I67, I68, I69 |  | 427.31, 427.32, 410, 411, 412, 413, 414, 430, 431, 432, 433, 434, 435, 436, 437, 438 | UKB variable 'Has a doctor ever told you that you have the following conditions 6150?'  1) Heart attack2) angina  3) stroke | 1471 atrial fibrillation  1483 atrial flutter  1074 angina  1075 heart attack/myocardial infarction  1081 stroke  1082 transient ischaemic attack 1083 subdural haemorrhage/haematoma  1086 subarachnoid haemorrhage |  |
| Ischaemic stroke | I63 |  | I48, I20, I21, I22, I23, I24, I25, G45, I60, I61, I62, I63, I64, I65, I66, I67, I68, I69 |  | 427.31, 427.32, 410, 411, 412, 413, 414, 430, 431, 432, 433, 434, 435, 436, 437, 438 | UKB variable 'Has a doctor ever told you that you have the following conditions 6150?'  1) Heart attack 2) angina,  3) stroke | 1471 atrial fibrillation  1483 atrial flutter  1074 angina  1075 heart attack/myocardial infarction  1081 stroke  1082 transient ischaemic attack1083 subdural haemorrhage/haematoma  1086 subarachnoid haemorrhage |  |
| Haemorrhagic stroke | I60 I61 |  | I48, I20, I21, I22, I23, I24, I25, G45, I60, I61, I62, I63, I64, I65, I66, I67, I68, I69 |  | 427.31, 427.32, 410, 411, 412, 413, 414, 430, 431, 432, 433, 434, 435, 436, 437, 438 | UKB variable 'Has a doctor ever told you that you have the following conditions 6150?'  1) Heart attack2) angina  3) stroke | 1471 atrial fibrillation  1483 atrial flutter  1074 angina  1075 heart attack/myocardial infarction  1081 stroke  1082 transient ischaemic attack 1083 subdural haemorrhage/haematoma  1086 subarachnoid haemorrhage |  |
| Venous thromboembolism | I26, I80, 181, I82 |  | I26, I80, 181, I82 |  | 415.1, 451, 452, 453 |  | 1068 venous thromboembolic disease  1094 deep venous thrombosis |  |
| Varicose veins of lower extremities | I83 | L84 L85 L86 L87 L88 | I83 | L84 L85 L86 L87 L88 | 454 |  | 1494 varicose veins | 1479 varicose vein surgery |
| Haemorrhoids | I84 | H51, H52, H53 | I84 | H51, H52, H53 | 455 |  | 1505 haemorrhoids / piles | 1483 haemorrhoidectomy / piles surgery/ banding of piles |
| Respiratory disease |  |  |  |  |  |  |  |  |
| Pneumonia | J18 |  | J18 |  | 480, 481, 482, 483, 484, 485, 486 |  | 1398 pneumonia |  |
| Digestive disease |  |  |  |  |  |  |  |  |
| Gastro-oesophageal reflux disease (GERD) | K21 |  | K21 |  | 530.11, 530.81 |  | 1138 gastro-oesophageal reflux (GERD) |  |
| Gastritis and duodenitis | K29 |  | K29 |  | 535 |  |  |  |
| Inguinal hernia | K40 | T19, T20, T21 | K40 | T19, T20, T21 | 550 |  | 1513 inguinal hernia | 1403 inguinal/femoral hernia repair  1563 inguinal hernia repair  1402 hernia surgery |
| Non-infective enteritis and colitis | K50, K51, K52 |  | K50, K51, K52 |  | 555, 556, 558 |  | 1459 colitis/not crohns or ulcerative colitis  1462 crohns disease  1463 ulcerative colitis |  |
| Diverticular disease of intestine | K57 |  | K57 |  | 562 |  | 1458 diverticular disease/diverticulitis |  |
| Colon polyps | D12, K63.5 | H20.1, H20.2, H20.3, H20.4, H20.5, H20.6, H22.1, H23.1,  H25.1, H28.1 | D12, K63.5 | H20.1, H20.2, H20.3, H20.4, H20.5, H20.6, H22.1, H23.1,  H25.1, H28.1 | 211.3, 211.4 |  | 1460 rectal or colon adenoma/polyps | 1463 rectal or colon polypectomy |
| Gallbladder disease | K80, K81 | J18 | K80, K81 | J18 | 574, 575.0, 575.1 |  | 1161 gall bladder disease | 1455 cholecystectomy/  gall bladder removal |
| Joint disorder |  |  |  |  |  |  |  |  |
| Osteoarthritis | M15, M16, M17, M18, M19, M47 |  | M15, M16, M17, M18, M19, M47 |  | 715, 721 |  | 1465 osteoarthritis |  |
| Genitourinary disease |  |  |  |  |  |  |  |  |
| Kidney stones | N20, N23 |  | N20, N23 |  | 592, 788.0 |  | 1197 kidney stone/ureter stone/bladder stone | 1197percutaneous/open kidney stone surgery/lithotripsy |
| Urinary tract infection | N39.0 |  | N39.0 |  | 599.0 |  | 1196 urinary tract infection/kidney infection |  |
| Hyperplasia of prostate | N40 |  | N40 |  | 600 |  | 1516 benign prostatic hypertrophy  1396 enlarged prostate |  |
| Female genital prolapse | N81 | M51, M52, M53, P22, P23, P24 | N81 | M51, M52, M53, P22, P23, P24 | 618 |  | 1353 vaginal prolapse/uterine prolapse | 1361 vaginal prolapse/  colposuspension |
| Other diseases |  |  |  |  |  |  |  |  |
| Uterine fibroids | D25 | Q10, Q18, Q074, Q089, Q171, Q092, Q093, Q161, T43 | D25 | Q10, Q18, Q074, Q089, Q171, Q092, Q093, Q161, T43 | 218 | UKB variable 'Ever had hysterectomy 3591'  1)yes  UKB variable 'Ever had menopause 2724'  2) don't know had hysterectomy | 1351 uterine fibroids | 1509 myomectomy/fibroids removed  1357 hysterectomy 1358 hysterectomy with oophorectomy 1359 hysterectomy with cervical sparing |
| Iron deficiency anaemia (IDA) | D50 |  | D50 |  | 280 |  | 1330 iron deficiency anaemia |  |
| Diabetes | E10, E11, E12, E13, E14 |  | E10, E11, E12, E13, E14 |  | 250 | UKB variable 'Dr. diagnosed diabetes 2443' 1)yes    UKB variable 'Taking medications 6177'  3) insulin | 1220 diabetes  1221 gestational diabetes  1222 type 1 diabetes  1223 type 2 diabetes |  |
| Carpal tunnel syndrome | G56.0 | A65.1 | G56.0 | A65.1 | 354.0 |  | 1541 carpal tunnel syndrome | 1501 carpal tunnel surgery |
| Cataract | H25, H26, Q120 | C71, C72, C73, C74, C75 | H25, H26, Q120, E10, E11, E13, E14 | C71, C72, C73, C74, C75 | 366, 250 | UKB variable 'Have eye problems 6148'  4) Cataract  UKB variable 'Dr. diagnosed diabetes 2443' 1)yes  UKB variable 'Taking medications 6177'  3) insulin | 1278 Cataract 1220 diabetes, 1221 gestational diabetes  1222 type 1 diabetes  1223 type 2 diabetes | 1435 cataract extraction/lens implant |
| Cellulitis | L03 |  | L03 |  | 681, 682 |  | 1625 cellulitis |  |
| ^a^Based on ICD-10 definition and UKB variable 20002 | | | | | | | | |
| ^b^Based on OPSC-4 definition and UKB variable 20004 | | | | | | | | |

| Supplementary Table S2. Relative risk of 25 common conditions per 5 nmol/l higher IGF-I concentration by various levels of adjustment. | | | | | | | | | |
| --- | --- | --- | --- | --- | --- | --- | --- | --- | --- |
|  | | **Insulin-like growth factor-I, sex-specific fifths** | | | | | | **Insulin-like growth factor-I**  **(per 5 nmol/l)** | |
| **Outcome** |  | **1st** | **2nd** | **3rd** | **4th** | **5th** | ***P*_trend_^a^** | uncorrected | corrected^b^ |
| Ischaemic heart disease | **Total n/cases** | 51792/2219 | 60177/2354 | 63003/2197 | 64101/2041 | 64688/1861 |  | 303761/10672 |  |
|  | HR (95%-CI), Model 0 | 1.00 (Ref) | 0.99 (0.93-1.05) | 0.95 (0.89-1.00) | 0.94 (0.88-1.00) | 0.97 (0.91-1.03) | 0.0636 | 0.98 (0.96-1.00) | 0.97 (0.95-1.00) |
|  | HR (95%-CI), Model 1 | 1.00 (Ref) | 1.01 (0.96-1.07) | 0.98 (0.92-1.04) | 0.98 (0.92-1.04) | 1.01 (0.95-1.08) | 0.8642 | 0.99 (0.98-1.01) | 0.99 (0.97-1.02) |
|  | HR (95%-CI), Model 2 | 1.00 (Ref) | 1.02 (0.97-1.09) | 0.99 (0.94-1.06) | 0.99 (0.94-1.06) | 1.03 (0.96-1.10) | 0.7720 | 1.00 (0.98-1.02) | 1.00 (0.98-1.02) |
|  | HR (95%-CI), Model 3 | 1.00 (Ref) | 1.03 (0.97-1.09) | 1.00 (0.95-1.07) | 1.01 (0.95-1.07) | 1.05 (0.98-1.12) | 0.3648 | 1.01 (0.99-1.03) | 1.01 (0.98-1.03) |
| Atrial fibrillation | **Total n/cases** | 51899/1113 | 60270/1110 | 63090/1004 | 64196/876 | 64749/759 |  | 304204/4862 |  |
|  | HR (95%-CI), Model 0 | 1.00 (Ref) | 0.95 (0.87-1.03) | 0.89 (0.82-0.97) | 0.84 (0.77-0.92) | 0.84 (0.77-0.93) | **<0.0001** | 0.94 (0.92-0.97) | 0.93 (0.90-0.96) |
|  | HR (95%-CI), Model 1 | 1.00 (Ref) | 0.94 (0.86-1.02) | 0.88 (0.81-0.96) | 0.83 (0.76-0.90) | 0.83 (0.75-0.91) | **<0.0001** | 0.94 (0.91-0.96) | 0.92 (0.89-0.96) |
|  | HR (95%-CI), Model 2 | 1.00 (Ref) | 0.97 (0.89-1.06) | 0.92 (0.84-1.00) | 0.87 (0.80-0.95) | 0.88 (0.80-0.97) | **0.0005** | 0.96 (0.93-0.99) | 0.95 (0.91-0.98) |
|  | HR (95%-CI), Model 3 | 1.00 (Ref) | 0.99 (0.91-1.08) | 0.95 (0.87-1.04) | 0.91 (0.83-1.00) | 0.93 (0.85-1.03) | 0.0330 | 0.98 (0.95-1.01) | 0.97 (0.94-1.01) |
| Cerebrovascular disease | **Total n/cases** | 51897/1243 | 60262/1179 | 63089/1087 | 64180/971 | 64765/851 |  | 304193/5331 |  |
|  | HR (95%-CI), Model 0 | 1.00 (Ref) | 0.92 (0.85-0.99) | 0.89 (0.82-0.97) | 0.87 (0.80-0.94) | 0.88 (0.80-0.96) | **0.0009** | 0.96 (0.94-0.99) | 0.95 (0.92-0.98) |
|  | HR (95%-CI), Model 1 | 1.00 (Ref) | 0.94 (0.87-1.02) | 0.92 (0.85-1.00) | 0.91 (0.83-0.99) | 0.92 (0.84-1.01) | 0.0397 | 0.98 (0.95-1.00) | 0.97 (0.94-1.00) |
|  | HR (95%-CI), Model 2 | 1.00 (Ref) | 0.95 (0.87-1.03) | 0.93 (0.86-1.01) | 0.92 (0.84-1.00) | 0.94 (0.85-1.02) | 0.0791 | 0.98 (0.96-1.01) | 0.98 (0.95-1.01) |
|  | HR (95%-CI), Model 3 | 1.00 (Ref) | 0.96 (0.88-1.04) | 0.95 (0.87-1.03) | 0.94 (0.86-1.03) | 0.97 (0.89-1.06) | 0.3914 | 0.99 (0.97-1.02) | 0.99 (0.96-1.03) |
| Ischaemic stroke | **Total n/cases** | 51928/598 | 60304/551 | 63121/505 | 64218/428 | 64793/397 |  | 304364/2479 |  |
|  | HR (95%-CI), Model 0 | 1.00 (Ref) | 0.90 (0.81-1.02) | 0.88 (0.78-0.99) | 0.83 (0.73-0.94) | 0.90 (0.79-1.02) | 0.0227 | 0.95 (0.92-0.99) | 0.94 (0.90-0.99) |
|  | HR (95%-CI), Model 1 | 1.00 (Ref) | 0.93 (0.83-1.04) | 0.92 (0.81-1.03) | 0.87 (0.76-0.98) | 0.95 (0.83-1.08) | 0.1761 | 0.97 (0.94-1.01) | 0.97 (0.92-1.01) |
|  | HR (95%-CI), Model 2 | 1.00 (Ref) | 0.94 (0.84-1.06) | 0.93 (0.83-1.05) | 0.89 (0.78-1.00) | 0.97 (0.85-1.11) | 0.3209 | 0.98 (0.94-1.02) | 0.98 (0.93-1.02) |
|  | HR (95%-CI), Model 3 | 1.00 (Ref) | 0.96 (0.85-1.08) | 0.96 (0.85-1.09) | 0.93 (0.81-1.05) | 1.03 (0.90-1.18) | 0.9970 | 1.00 (0.96-1.04) | 1.00 (0.95-1.05) |
| Haemorrhagic stroke | **Total n/cases** | 51942/218 | 60309/216 | 63125/210 | 64223/179 | 64796/158 |  | 304395/981 |  |
|  | HR (95%-CI), Model 0 | 1.00 (Ref) | 0.94 (0.78-1.14) | 0.95 (0.79-1.15) | 0.87 (0.71-1.07) | 0.87 (0.70-1.07) | 0.1250 | 0.96 (0.90-1.02) | 0.95 (0.88-1.03) |
|  | HR (95%-CI), Model 1 | 1.00 (Ref) | 0.98 (0.81-1.18) | 1.00 (0.83-1.22) | 0.93 (0.76-1.14) | 0.93 (0.76-1.15) | 0.4462 | 0.99 (0.93-1.05) | 0.98 (0.91-1.06) |
|  | HR (95%-CI), Model 2 | 1.00 (Ref) | 0.98 (0.81-1.18) | 1.00 (0.82-1.21) | 0.93 (0.76-1.13) | 0.93 (0.75-1.15) | 0.4207 | 0.98 (0.93-1.05) | 0.98 (0.91-1.06) |
|  | HR (95%-CI), Model 3 | 1.00 (Ref) | 0.99 (0.82-1.20) | 1.02 (0.84-1.24) | 0.95 (0.78-1.17) | 0.97 (0.78-1.20) | 0.6828 | 1.00 (0.94-1.06) | 1.00 (0.92-1.08) |
| Venous thromboembolism | **Total n/cases** | 54119/819 | 62263/734 | 64887/742 | 66015/706 | 66259/620 |  | 313543/3621 |  |
|  | HR (95%-CI), Model 0 | 1.00 (Ref) | 0.85 (0.77-0.93) | 0.88 (0.80-0.97) | 0.88 (0.80-0.98) | 0.86 (0.78-0.96) | 0.0314 | 0.96 (0.93-0.99) | 0.95 (0.91-0.99) |
|  | HR (95%-CI), Model 1 | 1.00 (Ref) | 0.85 (0.77-0.94) | 0.88 (0.80-0.98) | 0.89 (0.80-0.98) | 0.86 (0.77-0.96) | 0.0329 | 0.96 (0.93-0.99) | 0.95 (0.91-0.99) |
|  | HR (95%-CI), Model 2 | 1.00 (Ref) | 0.88 (0.79-0.97) | 0.92 (0.84-1.02) | 0.94 (0.84-1.04) | 0.91 (0.82-1.02) | 0.3175 | 0.98 (0.95-1.01) | 0.97 (0.94-1.01) |
|  | HR (95%-CI), Model 3 | 1.00 (Ref) | 0.89 (0.80-0.98) | 0.94 (0.85-1.04) | 0.96 (0.87-1.07) | 0.96 (0.86-1.07) | 0.9408 | 0.99 (0.96-1.03) | 0.99 (0.95-1.04) |
| Varicose veins | **Total n/cases** | 52106/417 | 59935/387 | 62655/394 | 63931/406 | 64504/335 |  | 303131/1939 |  |
|  | HR (95%-CI), Model 0 | 1.00 (Ref) | 0.83 (0.72-0.96) | 0.83 (0.72-0.95) | 0.86 (0.75-0.99) | 0.73 (0.63-0.84) | **0.0004** | 0.92 (0.88-0.96) | 0.89 (0.84-0.94) |
|  | HR (95%-CI), Model 1 | 1.00 (Ref) | 0.83 (0.73-0.96) | 0.83 (0.72-0.96) | 0.86 (0.75-0.99) | 0.72 (0.62-0.84) | **0.0004** | 0.91 (0.87-0.96) | 0.89 (0.83-0.94) |
|  | HR (95%-CI), Model 2 | 1.00 (Ref) | 0.85 (0.74-0.98) | 0.85 (0.74-0.98) | 0.88 (0.77-1.02) | 0.75 (0.64-0.87) | 0.0021 | 0.92 (0.88-0.97) | 0.90 (0.85-0.95) |
|  | HR (95%-CI), Model 3 | 1.00 (Ref) | 0.86 (0.75-0.99) | 0.87 (0.75-1.00) | 0.91 (0.79-1.05) | 0.78 (0.67-0.92) | 0.0198 | 0.94 (0.90-0.98) | 0.92 (0.86-0.98) |
| Haemorrhoids | **Total n/cases** | 53084/855 | 60872/1006 | 63320/1025 | 64373/1123 | 64683/1113 |  | 306332/5122 |  |
|  | HR (95%-CI), Model 0 | 1.00 (Ref) | 1.02 (0.93-1.12) | 1.00 (0.91-1.10) | 1.08 (0.99-1.18) | 1.07 (0.98-1.18) | 0.0555 | 1.03 (1.01-1.06) | 1.04 (1.01-1.07) |
|  | HR (95%-CI), Model 1 | 1.00 (Ref) | 1.03 (0.94-1.13) | 1.01 (0.93-1.11) | 1.10 (1.00-1.20) | 1.09 (0.99-1.20) | 0.0251 | 1.04 (1.01-1.06) | 1.05 (1.01-1.08) |
|  | HR (95%-CI), Model 2 | 1.00 (Ref) | 1.02 (0.93-1.12) | 1.00 (0.92-1.10) | 1.09 (0.99-1.19) | 1.07 (0.98-1.18) | 0.0525 | 1.03 (1.01-1.06) | 1.04 (1.01-1.08) |
|  | HR (95%-CI), Model 3 | 1.00 (Ref) | 1.02 (0.93-1.11) | 0.99 (0.90-1.09) | 1.07 (0.97-1.17) | 1.05 (0.95-1.15) | 0.1862 | 1.02 (1.00-1.05) | 1.03 (1.00-1.07) |
| Pneumonia | **Total n/cases** | 54436/1276 | 62424/1115 | 64966/1030 | 65949/891 | 66176/819 |  | 313951/5131 |  |
|  | HR (95%-CI), Model 0 | 1.00 (Ref) | 0.85 (0.79-0.92) | 0.83 (0.76-0.90) | 0.77 (0.71-0.84) | 0.81 (0.74-0.89) | **<0.0001** | 0.92 (0.90-0.95) | 0.90 (0.87-0.94) |
|  | HR (95%-CI), Model 1 | 1.00 (Ref) | 0.88 (0.81-0.95) | 0.87 (0.80-0.95) | 0.82 (0.75-0.90) | 0.87 (0.80-0.96) | **0.0003** | 0.95 (0.92-0.97) | 0.93 (0.90-0.97) |
|  | HR (95%-CI), Model 2 | 1.00 (Ref) | 0.90 (0.83-0.97) | 0.90 (0.83-0.98) | 0.85 (0.78-0.93) | 0.91 (0.83-1.00) | 0.0075 | 0.96 (0.93-0.99) | 0.95 (0.92-0.98) |
|  | HR (95%-CI), Model 3 | 1.00 (Ref) | 0.93 (0.86-1.01) | 0.95 (0.87-1.03) | 0.91 (0.83-1.00) | 1.00 (0.91-1.09) | 0.6233 | 0.99 (0.96-1.02) | 0.99 (0.96-1.02) |
| Gastro-oesophageal reflux disease (GERD) | **Total n/cases** | 52061/1083 | 60193/1155 | 62927/1154 | 64166/1177 | 64607/1149 |  | 303954/5718 |  |
|  | HR (95%-CI), Model 0 | 1.00 (Ref) | 0.96 (0.88-1.04) | 0.95 (0.87-1.03) | 0.98 (0.91-1.07) | 1.02 (0.93-1.11) | 0.5179 | 1.00 (0.98-1.02) | 1.00 (0.97-1.03) |
|  | HR (95%-CI), Model 1 | 1.00 (Ref) | 0.98 (0.90-1.06) | 0.97 (0.89-1.05) | 1.01 (0.93-1.10) | 1.05 (0.96-1.15) | 0.1585 | 1.01 (0.98-1.03) | 1.01 (0.98-1.04) |
|  | HR (95%-CI), Model 2 | 1.00 (Ref) | 0.98 (0.90-1.06) | 0.97 (0.89-1.05) | 1.01 (0.93-1.10) | 1.04 (0.96-1.14) | 0.2051 | 1.01 (0.98-1.03) | 1.01 (0.98-1.04) |
|  | HR (95%-CI), Model 3 | 1.00 (Ref) | 0.97 (0.89-1.06) | 0.96 (0.88-1.05) | 1.01 (0.92-1.10) | 1.04 (0.95-1.14) | 0.2305 | 1.01 (0.98-1.03) | 1.01 (0.98-1.04) |
| Gastritis and duodenitis | **Total n/cases** | 53396/1808 | 61551/1869 | 64256/1730 | 65350/1610 | 65818/1562 |  | 310371/8579 |  |
|  | HR (95%-CI), Model 0 | 1.00 (Ref) | 0.95 (0.89-1.01) | 0.87 (0.82-0.93) | 0.84 (0.78-0.90) | 0.87 (0.81-0.93) | **<0.0001** | 0.95 (0.93-0.97) | 0.94 (0.92-0.97) |
|  | HR (95%-CI), Model 1 | 1.00 (Ref) | 0.98 (0.92-1.05) | 0.92 (0.86-0.98) | 0.89 (0.83-0.95) | 0.93 (0.86-0.99) | **0.0012** | 0.97 (0.95-0.99) | 0.96 (0.94-0.99) |
|  | HR (95%-CI), Model 2 | 1.00 (Ref) | 0.98 (0.92-1.04) | 0.91 (0.86-0.98) | 0.88 (0.83-0.95) | 0.92 (0.86-0.99) | **0.0009** | 0.97 (0.95-0.99) | 0.96 (0.94-0.99) |
|  | HR (95%-CI), Model 3 | 1.00 (Ref) | 0.98 (0.91-1.04) | 0.91 (0.85-0.98) | 0.88 (0.82-0.94) | 0.92 (0.85-0.99) | **0.0007** | 0.97 (0.95-0.99) | 0.96 (0.94-0.99) |
| Inguinal hernia | **Total n/cases** | 51941/1301 | 59601/1532 | 62002/1517 | 63248/1463 | 63585/1352 |  | 300377/7165 |  |
|  | HR (95%-CI), Model 0 | 1.00 (Ref) | 1.10 (1.02-1.18) | 1.11 (1.03-1.20) | 1.11 (1.03-1.19) | 1.12 (1.03-1.21) | 0.0092 | 1.04 (1.02-1.06) | 1.05 (1.02-1.08) |
|  | HR (95%-CI), Model 1 | 1.00 (Ref) | 1.09 (1.01-1.17) | 1.10 (1.02-1.18) | 1.09 (1.01-1.17) | 1.09 (1.01-1.18) | 0.0544 | 1.03 (1.01-1.06) | 1.04 (1.01-1.07) |
|  | HR (95%-CI), Model 2 | 1.00 (Ref) | 1.05 (0.97-1.13) | 1.06 (0.98-1.14) | 1.04 (0.96-1.12) | 1.04 (0.96-1.12) | 0.4899 | 1.02 (0.99-1.04) | 1.02 (0.99-1.05) |
|  | HR (95%-CI), Model 3 | 1.00 (Ref) | 1.05 (0.97-1.13) | 1.05 (0.98-1.13) | 1.03 (0.95-1.11) | 1.03 (0.95-1.12) | 0.6595 | 1.01 (0.99-1.04) | 1.02 (0.99-1.05) |
| Non-infective enteritis and colitis | **Total n/cases** | 53978/875 | 61943/946 | 64456/878 | 65573/884 | 65729/809 |  | 311679/4392 |  |
|  | HR (95%-CI), Model 0 | 1.00 (Ref) | 0.97 (0.88-1.06) | 0.88 (0.80-0.97) | 0.90 (0.82-0.99) | 0.85 (0.77-0.94) | **0.0004** | 0.95 (0.93-0.98) | 0.94 (0.91-0.98) |
|  | HR (95%-CI), Model 1 | 1.00 (Ref) | 1.00 (0.91-1.10) | 0.92 (0.84-1.01) | 0.95 (0.86-1.04) | 0.91 (0.82-1.00) | 0.0252 | 0.97 (0.94-1.00) | 0.96 (0.93-1.00) |
|  | HR (95%-CI), Model 2 | 1.00 (Ref) | 1.00 (0.91-1.09) | 0.92 (0.84-1.01) | 0.95 (0.86-1.04) | 0.90 (0.82-1.00) | 0.0233 | 0.97 (0.94-1.00) | 0.96 (0.93-1.00) |
|  | HR (95%-CI), Model 3 | 1.00 (Ref) | 1.00 (0.91-1.09) | 0.92 (0.84-1.01) | 0.94 (0.86-1.04) | 0.90 (0.81-1.00) | 0.0233 | 0.97 (0.94-1.00) | 0.96 (0.93-1.00) |
| Diverticular disease | **Total n/cases** | 53804/2415 | 61861/2539 | 64532/2443 | 65647/2330 | 65994/2120 |  | 311838/11847 |  |
|  | HR (95%-CI), Model 0 | 1.00 (Ref) | 0.98 (0.92-1.03) | 0.95 (0.90-1.01) | 0.95 (0.89-1.00) | 0.94 (0.89-1.00) | 0.0262 | 0.99 (0.97-1.00) | 0.98 (0.96-1.00) |
|  | HR (95%-CI), Model 1 | 1.00 (Ref) | 0.99 (0.93-1.04) | 0.97 (0.92-1.03) | 0.97 (0.91-1.03) | 0.97 (0.92-1.03) | 0.2850 | 1.00 (0.98-1.01) | 1.00 (0.97-1.02) |
|  | HR (95%-CI), Model 2 | 1.00 (Ref) | 1.00 (0.94-1.06) | 0.98 (0.93-1.04) | 0.99 (0.93-1.05) | 0.99 (0.93-1.05) | 0.6470 | 1.00 (0.98-1.02) | 1.00 (0.98-1.02) |
|  | HR (95%-CI), Model 3 | 1.00 (Ref) | 1.00 (0.94-1.05) | 0.98 (0.93-1.04) | 0.99 (0.93-1.05) | 0.99 (0.93-1.05) | 0.6637 | 1.00 (0.98-1.02) | 1.00 (0.98-1.03) |
| Colon Polyps | **Total n/cases** | 52699/6171 | 60601/6640 | 63206/6566 | 64365/6390 | 64606/6085 |  | 305477/31852 |  |
|  | HR (95%-CI), Model 0 | 1.00 (Ref) | 0.97 (0.94-1.00) | 0.95 (0.91-0.98) | 0.94 (0.91-0.97) | 0.94 (0.91-0.98) | **0.0004** | 0.99 (0.98-1.00) | 0.98 (0.97-1.00) |
|  | HR (95%-CI), Model 1 | 1.00 (Ref) | 0.98 (0.95-1.02) | 0.97 (0.93-1.00) | 0.96 (0.93-1.00) | 0.98 (0.94-1.01) | 0.1228 | 1.00 (0.99-1.01) | 1.00 (0.98-1.01) |
|  | HR (95%-CI), Model 2 | 1.00 (Ref) | 0.99 (0.95-1.02) | 0.97 (0.94-1.01) | 0.97 (0.94-1.01) | 0.99 (0.95-1.02) | 0.3112 | 1.00 (0.99-1.01) | 1.00 (0.99-1.01) |
|  | HR (95%-CI), Model 3 | 1.00 (Ref) | 0.99 (0.95-1.02) | 0.97 (0.94-1.01) | 0.97 (0.93-1.01) | 0.98 (0.95-1.02) | 0.2147 | 1.00 (0.99-1.01) | 1.00 (0.99-1.01) |
| Gallbladder disease | **Total n/cases** | 52741/1414 | 60971/1421 | 63756/1349 | 64955/1278 | 65303/1266 |  | 307726/6728 |  |
|  | HR (95%-CI), Model 0 | 1.00 (Ref) | 0.89 (0.83-0.96) | 0.83 (0.77-0.90) | 0.79 (0.73-0.85) | 0.81 (0.75-0.88) | **<0.0001** | 0.94 (0.92-0.96) | 0.92 (0.89-0.95) |
|  | HR (95%-CI), Model 1 | 1.00 (Ref) | 0.92 (0.85-0.99) | 0.86 (0.80-0.93) | 0.82 (0.76-0.88) | 0.83 (0.77-0.90) | **<0.0001** | 0.95 (0.92-0.97) | 0.93 (0.91-0.96) |
|  | HR (95%-CI), Model 2 | 1.00 (Ref) | 0.97 (0.90-1.04) | 0.92 (0.86-1.00) | 0.90 (0.83-0.97) | 0.92 (0.85-1.00) | 0.0088 | 0.98 (0.96-1.00) | 0.97 (0.94-1.00) |
|  | HR (95%-CI), Model 3 | 1.00 (Ref) | 0.97 (0.90-1.05) | 0.94 (0.87-1.01) | 0.92 (0.85-1.00) | 0.96 (0.89-1.05) | 0.1676 | 0.99 (0.97-1.02) | 0.99 (0.96-1.02) |
| Osteoarthritis | **Total n/cases** | 49327/3579 | 57416/3623 | 60430/3531 | 61876/3205 | 62793/2945 |  | 291842/16883 |  |
|  | HR (95%-CI), Model 0 | 1.00 (Ref) | 0.95 (0.90-0.99) | 0.95 (0.90-0.99) | 0.91 (0.87-0.95) | 0.94 (0.90-0.99) | 0.0025 | 0.98 (0.97-1.00) | 0.98 (0.96-0.99) |
|  | HR (95%-CI), Model 1 | 1.00 (Ref) | 0.96 (0.92-1.01) | 0.96 (0.92-1.01) | 0.93 (0.89-0.98) | 0.97 (0.93-1.02) | 0.1163 | 0.99 (0.98-1.01) | 0.99 (0.97-1.01) |
|  | HR (95%-CI), Model 2 | 1.00 (Ref) | 1.00 (0.96-1.05) | 1.02 (0.97-1.07) | 1.00 (0.95-1.05) | 1.05 (1.00-1.11) | 0.1027 | 1.02 (1.00-1.03) | 1.02 (1.00-1.04) |
|  | HR (95%-CI), Model 3 | 1.00 (Ref) | 1.01 (0.96-1.06) | 1.03 (0.98-1.08) | 1.01 (0.97-1.07) | 1.08 (1.02-1.13) | 0.0134 | 1.02 (1.01-1.04) | 1.03 (1.01-1.05) |
| Kidney stones | **Total n/cases** | 54655/339 | 62607/324 | 65090/387 | 66057/372 | 66252/362 |  | 314661/1784 |  |
|  | HR (95%-CI), Model 0 | 1.00 (Ref) | 0.82 (0.71-0.96) | 0.94 (0.82-1.09) | 0.89 (0.77-1.04) | 0.87 (0.75-1.02) | 0.2887 | 0.99 (0.95-1.04) | 0.99 (0.94-1.05) |
|  | HR (95%-CI), Model 1 | 1.00 (Ref) | 0.84 (0.72-0.98) | 0.97 (0.83-1.12) | 0.91 (0.79-1.06) | 0.88 (0.76-1.03) | 0.3828 | 1.00 (0.95-1.04) | 1.00 (0.94-1.05) |
|  | HR (95%-CI), Model 2 | 1.00 (Ref) | 0.86 (0.74-1.00) | 1.00 (0.86-1.16) | 0.95 (0.82-1.11) | 0.92 (0.79-1.08) | 0.7662 | 1.01 (0.97-1.06) | 1.01 (0.96-1.07) |
|  | HR (95%-CI), Model 3 | 1.00 (Ref) | 0.84 (0.72-0.98) | 0.97 (0.83-1.12) | 0.91 (0.78-1.06) | 0.87 (0.74-1.02) | 0.2751 | 0.99 (0.95-1.04) | 0.99 (0.93-1.05) |
| Urinary tract infection | **Total n/cases** | 54627/866 | 62585/819 | 65080/792 | 66085/748 | 66220/675 |  | 314597/3900 |  |
|  | HR (95%-CI), Model 0 | 1.00 (Ref) | 0.92 (0.84-1.02) | 0.94 (0.85-1.03) | 0.95 (0.86-1.04) | 0.97 (0.87-1.07) | 0.6615 | 0.99 (0.96-1.02) | 0.98 (0.95-1.02) |
|  | HR (95%-CI), Model 1 | 1.00 (Ref) | 0.95 (0.86-1.05) | 0.98 (0.89-1.08) | 0.99 (0.90-1.10) | 1.01 (0.91-1.12) | 0.6078 | 1.00 (0.97-1.03) | 1.00 (0.96-1.04) |
|  | HR (95%-CI), Model 2 | 1.00 (Ref) | 0.97 (0.88-1.06) | 1.00 (0.91-1.10) | 1.02 (0.92-1.13) | 1.04 (0.94-1.16) | 0.2667 | 1.01 (0.98-1.04) | 1.01 (0.98-1.05) |
|  | HR (95%-CI), Model 3 | 1.00 (Ref) | 0.98 (0.89-1.08) | 1.03 (0.93-1.13) | 1.06 (0.96-1.17) | 1.10 (0.98-1.22) | 0.0407 | 1.03 (1.00-1.06) | 1.04 (1.00-1.08) |
| Hyperplasia of prostate | **Total n/cases** | 23141/573 | 26879/664 | 27917/664 | 28426/620 | 28315/579 |  | 134678/3100 |  |
|  | HR (95%-CI), Model 0 | 1.00 (Ref) | 1.11 (1.00-1.25) | 1.17 (1.05-1.31) | 1.17 (1.04-1.31) | 1.25 (1.11-1.41) | **0.0002** | 1.07 (1.03-1.10) | 1.08 (1.04-1.13) |
|  | HR (95%-CI), Model 1 | 1.00 (Ref) | 1.10 (0.98-1.23) | 1.15 (1.03-1.28) | 1.13 (1.01-1.27) | 1.20 (1.06-1.35) | 0.0038 | 1.05 (1.02-1.09) | 1.07 (1.02-1.11) |
|  | HR (95%-CI), Model 2 | 1.00 (Ref) | 1.09 (0.98-1.22) | 1.14 (1.02-1.27) | 1.12 (1.00-1.26) | 1.18 (1.05-1.33) | 0.0079 | 1.05 (1.01-1.08) | 1.06 (1.02-1.11) |
|  | HR (95%-CI), Model 3 | 1.00 (Ref) | 1.08 (0.97-1.21) | 1.12 (1.00-1.26) | 1.10 (0.98-1.24) | 1.16 (1.03-1.31) | 0.0236 | 1.04 (1.01-1.08) | 1.05 (1.01-1.10) |
| Female genital prolapse | **Total n/cases** | 29138/895 | 33476/886 | 35114/955 | 35787/847 | 36331/879 |  | 169846/4462 |  |
|  | HR (95%-CI), Model 0 | 1.00 (Ref) | 0.90 (0.82-0.99) | 0.97 (0.88-1.06) | 0.89 (0.81-0.97) | 0.99 (0.90-1.09) | 0.6674 | 1.00 (0.97-1.03) | 1.00 (0.96-1.04) |
|  | HR (95%-CI), Model 1 | 1.00 (Ref) | 0.93 (0.85-1.02) | 1.02 (0.93-1.12) | 0.93 (0.85-1.03) | 1.05 (0.96-1.16) | 0.3510 | 1.02 (0.99-1.05) | 1.03 (0.99-1.07) |
|  | HR (95%-CI), Model 2 | 1.00 (Ref) | 0.93 (0.84-1.02) | 1.01 (0.92-1.11) | 0.93 (0.84-1.02) | 1.04 (0.94-1.15) | 0.4619 | 1.02 (0.99-1.05) | 1.02 (0.99-1.06) |
|  | HR (95%-CI), Model 3 | 1.00 (Ref) | 0.93 (0.84-1.02) | 1.02 (0.92-1.11) | 0.93 (0.84-1.03) | 1.04 (0.94-1.16) | 0.4277 | 1.02 (0.99-1.05) | 1.03 (0.99-1.07) |
| Uterine fibroids | **Total n/cases** | 20355/1056 | 25503/1291 | 27629/1435 | 28960/1556 | 29989/1720 |  | 132436/7058 |  |
|  | HR (95%-CI), Model 0 | 1.00 (Ref) | 0.92 (0.85-1.00) | 0.91 (0.84-0.99) | 0.90 (0.83-0.97) | 0.89 (0.82-0.96) | 0.0073 | 0.97 (0.95-0.99) | 0.96 (0.93-0.99) |
|  | HR (95%-CI), Model 1 | 1.00 (Ref) | 0.94 (0.87-1.02) | 0.93 (0.86-1.01) | 0.92 (0.85-1.00) | 0.92 (0.85-0.99) | 0.0540 | 0.98 (0.96-1.00) | 0.97 (0.94-1.00) |
|  | HR (95%-CI), Model 2 | 1.00 (Ref) | 0.97 (0.89-1.05) | 0.98 (0.90-1.06) | 0.98 (0.90-1.06) | 0.99 (0.91-1.07) | 0.9639 | 1.00 (0.98-1.02) | 1.00 (0.97-1.03) |
|  | HR (95%-CI), Model 3 | 1.00 (Ref) | 0.97 (0.89-1.05) | 0.98 (0.90-1.06) | 0.98 (0.90-1.06) | 0.99 (0.91-1.07) | 0.9584 | 1.00 (0.97-1.02) | 1.00 (0.96-1.03) |
| Iron deficiency anaemia | **Total n/cases** | 54790/882 | 62792/802 | 65297/759 | 66346/686 | 66431/637 |  | 315656/3766 |  |
|  | HR (95%-CI), Model 0 | 1.00 (Ref) | 0.84 (0.76-0.92) | 0.80 (0.72-0.88) | 0.74 (0.67-0.82) | 0.73 (0.65-0.81) | **<0.0001** | 0.90 (0.87-0.93) | 0.88 (0.84-0.91) |
|  | HR (95%-CI), Model 1 | 1.00 (Ref) | 0.86 (0.78-0.95) | 0.82 (0.75-0.91) | 0.77 (0.69-0.85) | 0.75 (0.67-0.83) | **<0.0001** | 0.91 (0.88-0.94) | 0.89 (0.85-0.92) |
|  | HR (95%-CI), Model 2 | 1.00 (Ref) | 0.87 (0.79-0.96) | 0.84 (0.76-0.93) | 0.78 (0.71-0.87) | 0.77 (0.69-0.85) | **<0.0001** | 0.92 (0.89-0.95) | 0.90 (0.86-0.93) |
|  | HR (95%-CI), Model 3 | 1.00 (Ref) | 0.89 (0.81-0.98) | 0.87 (0.78-0.96) | 0.82 (0.74-0.91) | 0.81 (0.73-0.91) | **<0.0001** | 0.94 (0.91-0.97) | 0.92 (0.88-0.96) |
| Diabetes | **Total n/cases** | 55394/2217 | 63412/1645 | 65929/1518 | 66924/1328 | 67090/1237 |  | 318749/7945 |  |
|  | HR (95%-CI), Model 0 | 1.00 (Ref) | 0.70 (0.65-0.74) | 0.66 (0.62-0.70) | 0.61 (0.57-0.65) | 0.63 (0.59-0.68) | **<0.0001** | 0.84 (0.82-0.86) | 0.80 (0.78-0.82) |
|  | HR (95%-CI), Model 1 | 1.00 (Ref) | 0.74 (0.69-0.78) | 0.71 (0.67-0.76) | 0.67 (0.62-0.72) | 0.70 (0.65-0.75) | **<0.0001** | 0.87 (0.85-0.89) | 0.84 (0.82-0.87) |
|  | HR (95%-CI), Model 2 | 1.00 (Ref) | 0.81 (0.76-0.87) | 0.82 (0.76-0.87) | 0.79 (0.74-0.85) | 0.85 (0.79-0.91) | **<0.0001** | 0.94 (0.92-0.96) | 0.92 (0.90-0.95) |
|  | HR (95%-CI), Model 3 | 1.00 (Ref) | 0.81 (0.76-0.86) | 0.80 (0.75-0.86) | 0.77 (0.72-0.83) | 0.80 (0.75-0.87) | **<0.0001** | 0.92 (0.90-0.94) | 0.90 (0.88-0.93) |
| Carpal tunnel syndrome | **Total n/cases** | 54415/624 | 62384/766 | 64940/697 | 65861/728 | 66078/715 |  | 313678/3530 |  |
|  | HR (95%-CI), Model 0 | 1.00 (Ref) | 1.13 (1.01-1.25) | 1.02 (0.91-1.13) | 1.08 (0.97-1.21) | 1.12 (1.00-1.25) | 0.1570 | 1.04 (1.00-1.07) | 1.05 (1.01-1.09) |
|  | HR (95%-CI), Model 1 | 1.00 (Ref) | 1.19 (1.07-1.32) | 1.09 (0.98-1.22) | 1.19 (1.07-1.32) | 1.25 (1.12-1.40) | **0.0006** | 1.07 (1.04-1.10) | 1.09 (1.05-1.13) |
|  | HR (95%-CI), Model 2 | 1.00 (Ref) | 1.23 (1.11-1.37) | 1.15 (1.03-1.28) | 1.27 (1.13-1.41) | 1.34 (1.20-1.50) | **<0.0001** | 1.09 (1.06-1.13) | 1.12 (1.08-1.16) |
|  | HR (95%-CI), Model 3 | 1.00 (Ref) | 1.22 (1.10-1.36) | 1.13 (1.02-1.27) | 1.24 (1.11-1.38) | 1.30 (1.15-1.46) | **0.0001** | 1.08 (1.05-1.12) | 1.10 (1.06-1.15) |
| Cataracts | **Total n/cases** | 52695/5206 | 60964/4749 | 63735/4387 | 64887/3894 | 65449/3054 |  | 307730/21290 |  |
|  | HR (95%-CI), Model 0 | 1.00 (Ref) | 0.91 (0.88-0.95) | 0.91 (0.88-0.95) | 0.91 (0.87-0.95) | 0.88 (0.84-0.92) | **<0.0001** | 0.97 (0.95-0.98) | 0.96 (0.94-0.97) |
|  | HR (95%-CI), Model 1 | 1.00 (Ref) | 0.92 (0.89-0.96) | 0.93 (0.89-0.97) | 0.93 (0.89-0.97) | 0.90 (0.86-0.94) | **<0.0001** | 0.97 (0.96-0.99) | 0.97 (0.95-0.98) |
|  | HR (95%-CI), Model 2 | 1.00 (Ref) | 0.93 (0.89-0.96) | 0.93 (0.89-0.97) | 0.94 (0.90-0.98) | 0.91 (0.87-0.95) | **0.0002** | 0.98 (0.96-0.99) | 0.97 (0.95-0.99) |
|  | HR (95%-CI), Model 3 | 1.00 (Ref) | 0.93 (0.89-0.97) | 0.94 (0.90-0.98) | 0.94 (0.90-0.99) | 0.92 (0.88-0.96) | 0.0022 | 0.98 (0.97-0.99) | 0.97 (0.96-0.99) |
| Cellulitis | **Total n/cases** | 55001/597 | 63021/503 | 65570/515 | 66572/462 | 66720/478 |  | 316884/2555 |  |
|  | HR (95%-CI), Model 0 | 1.00 (Ref) | 0.78 (0.69-0.87) | 0.80 (0.71-0.90) | 0.73 (0.65-0.83) | 0.80 (0.71-0.91) | **0.0002** | 0.93 (0.89-0.96) | 0.91 (0.86-0.95) |
|  | HR (95%-CI), Model 1 | 1.00 (Ref) | 0.78 (0.70-0.88) | 0.81 (0.72-0.91) | 0.75 (0.66-0.85) | 0.82 (0.72-0.93) | 0.0014 | 0.93 (0.90-0.97) | 0.92 (0.87-0.96) |
|  | HR (95%-CI), Model 2 | 1.00 (Ref) | 0.84 (0.75-0.95) | 0.89 (0.79-1.00) | 0.84 (0.74-0.95) | 0.93 (0.82-1.06) | 0.2759 | 0.98 (0.94-1.01) | 0.97 (0.92-1.02) |
|  | HR (95%-CI), Model 3 | 1.00 (Ref) | 0.86 (0.76-0.97) | 0.92 (0.82-1.04) | 0.88 (0.77-1.00) | 1.00 (0.87-1.13) | 0.9535 | 1.00 (0.96-1.03) | 0.99 (0.95-1.04) |

**^a^***P*trend bold: *P*<0.002. ^b^Corrected for regression dilution bias

**Model 0**: Stratified for age group (<45, 45–49, 50–54, 55–59, 60–64, and ≥65 years), sex and region and adjusted for age (underlying time variable). **Model 1**: Model 0 additionally adjusted for ethnicity (White, non-white, unknown), deprivation (Townsend index quintiles, unknown), qualification (College or university degree/vocational qualification, National examination at ages 17-18,National examination at age 16, unknown), smoking (never, former, current <15 cigarettes/day, current >15 cigarettes/ day, unknown), physical activity (<10, 10-<20, 20-40, 40-<60, >=60 MET hours per week, unknown), alcohol intake (<1.0, 1.0–4.9, 5.0–9.9, 10.0–14.9, 15.0–19.9, 20.0–24.9, and ≥25.0 g/day, non-drinker, and unknown), height (continuous), and in women: menopausal status (pre-, postmenopausal, unknown), hormone-replacement therapy (never, past, current, unknown), oral contraceptive pill intake (never, past, current, unknown), and parity (nulliparous, 1-2, 3 or more, unknown). **Model 2** (main model): Model 1 additionally adjusted for body mass index (<20.0, 20.0-22.49, 22.5-24.99, 25.0-27.49, 27.5-29.99, 30.0-32.49, 32.5-34.99, >35.0). **Model 3** (sensitivity analysis): Model 2 additionally adjusted for serum concentrations of C-reactive protein (quintiles, unknown), glycated haemoglobin (quintiles, unknown), and sex hormone-binding globulin (quintiles, unknown). CI confidence intervals.

### Supplementary Figures

503 317 participants recruited (2006-2010)

Basic exclusion criteria:

- Withdrawn consent (n=829)
- No biomarker data (n=4066)
- No IGF-I data at baseline (n=31 397)

467 025 participants

Health related exclusion criteria:

- Genetic sex differed to reported gender (n=355)
- Missing information for weight (n=1553)
- Missing information for height (n=307)
- Had prevalent cancer (n=24 626)
- Not in good or excellent health (n=110 830)
- Had unknown or prevalent diabetes (n=10 425)
- Had less than 1 year of follow-up (n=180)

**318 749 maximal analytical sample**

#### Supplementary Figure S1. Participant flow chart.
